# Consistency and applicability of return to activity guidelines in tactical-athletes with exertional heat illness. A systematic review

**DOI:** 10.1101/2022.08.03.22278365

**Authors:** Matthew O’Reilly, Yao-Wen Eliot Hu, Jonathan Gruber, Douglas M. Jones, Arthur Daniel, Janelle Marra, John J. Fraser

## Abstract

**Objective:** To assess the consistency of return to sport and occupation recommendations following EHI provided in published clinical practice guidelines, consensus statements, position statements, and practice alerts. A secondary aim was to evaluate the consistency of medical policies governing the return to duty following EHI between the branches of the United States Armed Forces and the agreement with published recommendations.

**Methods:** Ovid MEDLINE, Web of Science, and CINAHL databases were searched for clinical practice guidelines and position statements published at any time that guided return to activity in individuals with EHI. Methodological quality was assessed and the specific recommendations for clinical management were extracted. Consistency of recommendations was evaluated.

Agreement between published guidelines and the policies governing return to activity in military tactical athletes with heat injury were also evaluated.

**Results:** Two professional societal guidelines provided recommendations pertaining to return to function following EHI. There was consistency between guidelines regarding recommendations that addressed abstinence from activity; medical follow-up; graded resumption of activity; and return to function. Pertaining to military policy, contemporary regulations published in recent years reflected the best evidence provided in the professional guidelines. The greatest incongruency was noted in older military policies.

**Conclusions:** This systematic review highlights the need for consistent recommendation across all branches of the military when it comes to returning servicemembers to duty after EHI.

## INTRODUCTION

Exertional heat illness (EHI) is a clinical condition that encompasses both heat exhaustion and exertional heat stroke.^25^ Heat exhaustion is defined as the inability to continue exercising with a core body temperature less than 40°C (typically ranging between 38.5°C and 40°C), with nausea, vomiting, headache, fainting, weakness and cold or clammy skin common.^25,26^ Exertional heat stroke is substantially more severe, involves core temperatures greater than 40°C, and typically results in multiorgan failure and central nervous system dysfunction.^25,26^ These conditions, which frequently affect athletes engaged in sport and tactical occupations, occur when physiological thermoregulation is impaired or cannot maintain equilibrium with heat generated during metabolism or environmental exposure.^25^ Sports-related EHI occurs up to 4.19 per 1000 athlete-exposures during participation in American football.^13^ The prevalence of EHI during more extreme sports during competition in warm environments is even more common, with up to 54.5% of desert ultramarathoners found to have EHI.^13^ In military servicemembers (commonly referred to as tactical-athletes), these conditions occur at a rate of 0.48 per 1000 person-years.^1^ With climbing environmental temperatures expected with the changing climate, EHI is expected to become more problematic in the future.^10^

As a result of common sequelae of EHI, which includes reduced exercise capacity and heat intolerance,^22^ substantial limitations in physical activity and participation in sport or occupational tasks are likely. As such, impairments and activity limitations resulting from EHI not only affect the health of the individual tactical athlete, but can also have a profound effect on the ability of an organization (i.e. team or unit) to meet operational objectives. Therefore, proper clinical management is especially salient to organizational function, especially when lives are dependent on the successful execution of the mission.

The synthesis of findings and recommendations presented in clinical practice guidelines (CPGs), consensus statements, position statements, and practice alerts are highly useful during evidence-based decision-making, especially when planning resumption of sport and occupational activities following injury and illness. While CPGs have been found to be effective in improving the process and structure of medical care, they are frequently heterogenous in their recommendations.^17^ Similarly, resumption of activity may be dictated by organizational policy informed by medical evidence and balanced by operational demands and risk management decision-making, factors that are also likely to be heterogenous. To our knowledge, there are limited professional society recommendations or consensus statements for progressive return to activity after EHI in the current literature and there have been no systematic reviews that have evaluated the consistency of recommendations pertaining to return to sport or occupation following EHI. Furthermore, an evaluation of the medical policies of the United States Armed Forces governing the return to duty following EHI and the congruency of the policies with published guidelines is also warranted to ensure that the most updated evidence-based management is provided to military tactical-athletes. Therefore, the purpose of this systematic review was to assess the consistency of return to sport and occupation recommendations following EHI provided in published CPGs, consensus statements, position statements, and practice alerts. A secondary aim was to evaluate the consistency of medical policies governing the return to duty following EHI between the branches of the United States Armed Forces and the agreement with published recommendations.

## METHODS

The protocol for this study was registered *a priori* in PROSPERO (CRD42020216532, http://bit.ly/MilitaryHeatRTD). The Preferred Reporting Items for Systematic Reviews and Meta-Analyses (PRISMA)^24^ and A MeaSurement Tool to Assess systematic Reviews version 2 (AMSTAR 2)^29^ were used to guide study reporting.

### Eligibility Criteria

Guidelines were eligible for inclusion if they were a consensus guideline, society guideline, or CPG published at any time and addressed resumption of physical activity or duty requirements following EHI in adults.

### Search Strategy

A research librarian was consulted to develop the search strategy. The search strategy, comprised of MeSH terms, subjects, and keywords is detailed in **Supplemental Table 1**. The searches were limited to records in English, the native language of the study team, published at any time of inquiry. Ovid MEDLINE, Web of Science (including Science Citation Index Expanded and Social Sciences Citation Index), and CINAHL, were queried on 31 December 2019 and again on 31 January 2022 for CPGs, consensus statements, position statements, and practice alerts published at any time. and duplicates were removed using EndNote deduplication.^4^ Records were organized, reviewed, and selected using Rayyan QCRI, an application used to facilitate study selection for systematic reviews (https://rayyan.qcri.org/).

Two reviewers (MO and JG) independently reviewed each record by title and then abstract for inclusion. A third author (DJ) resolved any disagreements. Study selection is detailed in the PRISMA flowsheet (Figure 1).

**Figure 1.**
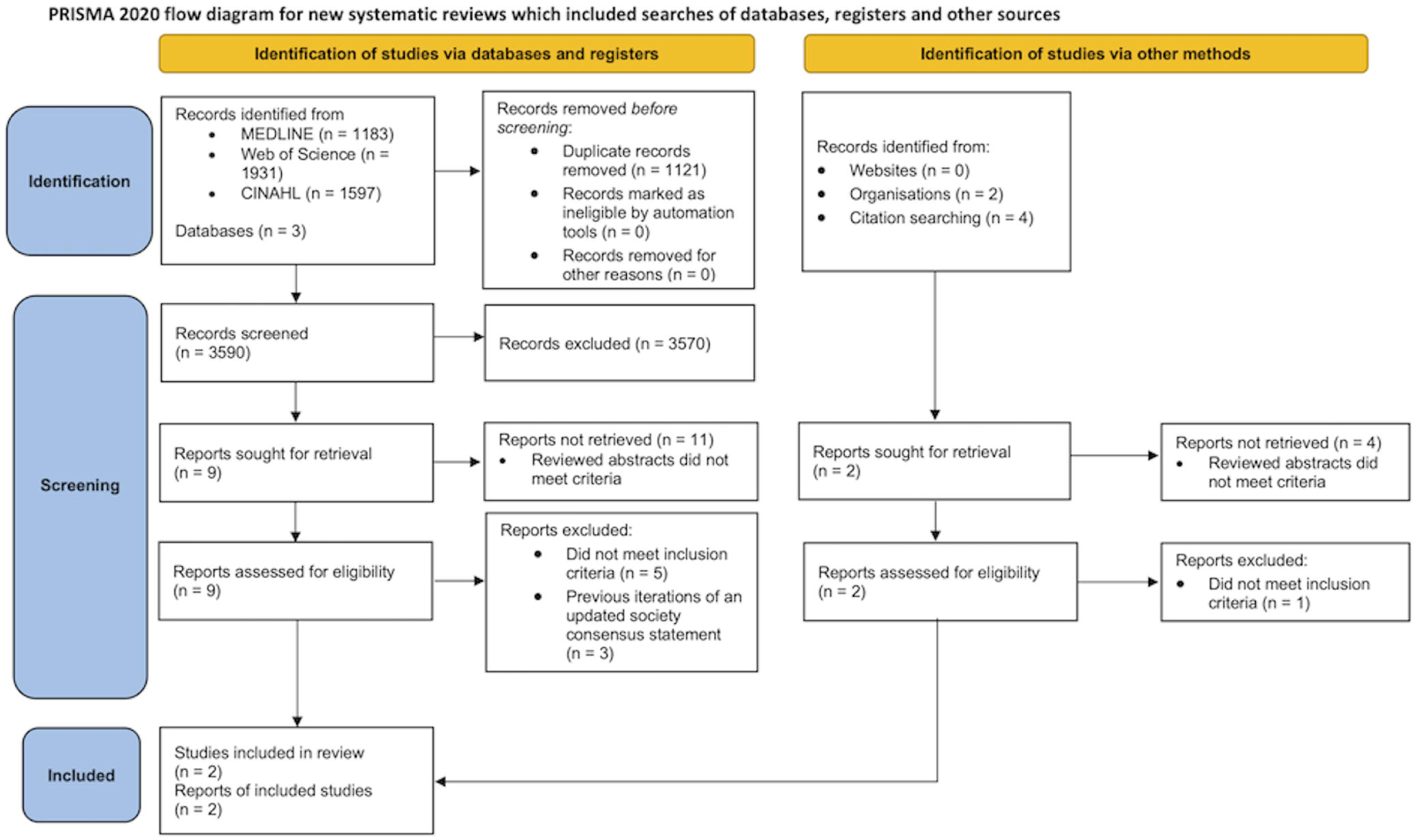
PRISMA flowsheet

### Data Extraction

Two reviewers (MO and JG) independently assessed each guideline and extracted the specific recommendations for return to activity. Characteristics of the clinical guidelines are reported in **Supplemental table 2**. Similarly, policy statements guiding the return to duty in the United States Armed Forces were extracted using the same approach. Any disagreements were resolved by consensus. If consensus could not be achieved, a third author (DJ) resolved any disagreements.

### Risk of Bias Assessment

Methodological quality of the included guidelines were assessed using the AGREE II tool.^5^ While there are acknowledged differences in methodological quality between CPGs and consensus statements,^18^ the AGREE II was employed in the current study to assess methodological quality across the full spectrum of guidelines. Each study was assessed in seven domains: Scope and Purpose; Stakeholder Involvement; Rigor of Development; Clarity of Presentation; Applicability; and Editorial Independence. Each domain was independently rated by three authors (JG, AD, and DJ). Reviewers resolved disagreements by consensus, and a fourth author (YEH) was consulted to resolve disagreements if needed.

### Synthesis Methods

Consistency of recommendations between published guidelines and congruency of military medical policy with practice guidelines was evaluated using a matrix. Based on the nature of the current study and the limited number of guidelines addressing this topic, a meta-analysis was not performed.

## RESULTS

### Selection

The search strategy yielded 3,590 unique records after duplicates were removed, with four records identified through citation cross-referencing (**Figure 1**). The authors also searched content from the major medical associations representing sports medicine, family medicine, emergency medicine, occupational medicine, and wilderness medicine, which yielded two additional guidelines for review.^14,15^ Based on the 24 abstracts reviewed for appropriateness, 13 abstracts failed to meet criteria for this study since they did not address return to sport or activity following EHI. Of the 11 full-text records assessed for eligibility,^2,3,7–9,14,15,22,23,25,26^ three records were excluded^2,3,8^ since they were previous versions of more recent society consensus statements.^26^ Of the remaining records, six were excluded^7,14,15,22,23,25^ as they were not a consensus statement, CPG, society guideline, or did not specifically provide recommendations for resumption of activity following EHI. Two records were included in the systematic review.^9,26^

Pertaining to military medical policy in the US Armed Forces, the following regulations were identified and reviewed for specific guidance pertaining to return to duty following EHI: Department of Defense Medical Standards for Military Service, Retention (DOD Instruction 6130.03, Volume 2); Navy Environmental Health Center Prevention of Heat/Cold Injuries (NEHC-TM-OEM 6260.6A); Manual of Naval Preventive Medicine Chapter 3, Prevention of Heat and Cold Stress Injuries (NAVMED P-5010-3); Marine Corps Heat Injury Prevention Program (Marine Corps Order 6200.1E w/Ch 1); US Army Standards of Medical Fitness (Army Regulation 40–501); Army & Air Force Heat Stress Control and Heat Casualty Management (TB MED 507/AFPAM 48-152); and US Army Training and Doctrine Command Prevention of Heat And Cold Casualties (TRADOC Regulation 350-29). Of these policy statements, only NEHC-TM-OEM 6260.6A, AR 40–501, and TB MED 507 provided specific clinical guidance pertaining to return to activity following EHI.

### Study Characteristics

**Table 1** details the extracted recommendations from the included societal guidelines and military policies. The included records consisted of a consensus statement from the American College of Sports Medicine (ACSM)^26^ and a position statement from the National Athletic Trainers’ Association (NATA).^9^ While a brief report by O’Connor, et al.^23^ summarizing the US military health policy pertaining to return to activity following EHI did not meet inclusion criteria as a guideline, this work provided context to how policy has changed since 2007 and was reported in **Table** 1. Both the ACSM and NATA statements provided practice recommendations regarding clinical management of EHI, to include prevention, recognition, treatment, and progressive resumption of physical activity in the general population and athletes alike.^9,26^

**Table 1.**
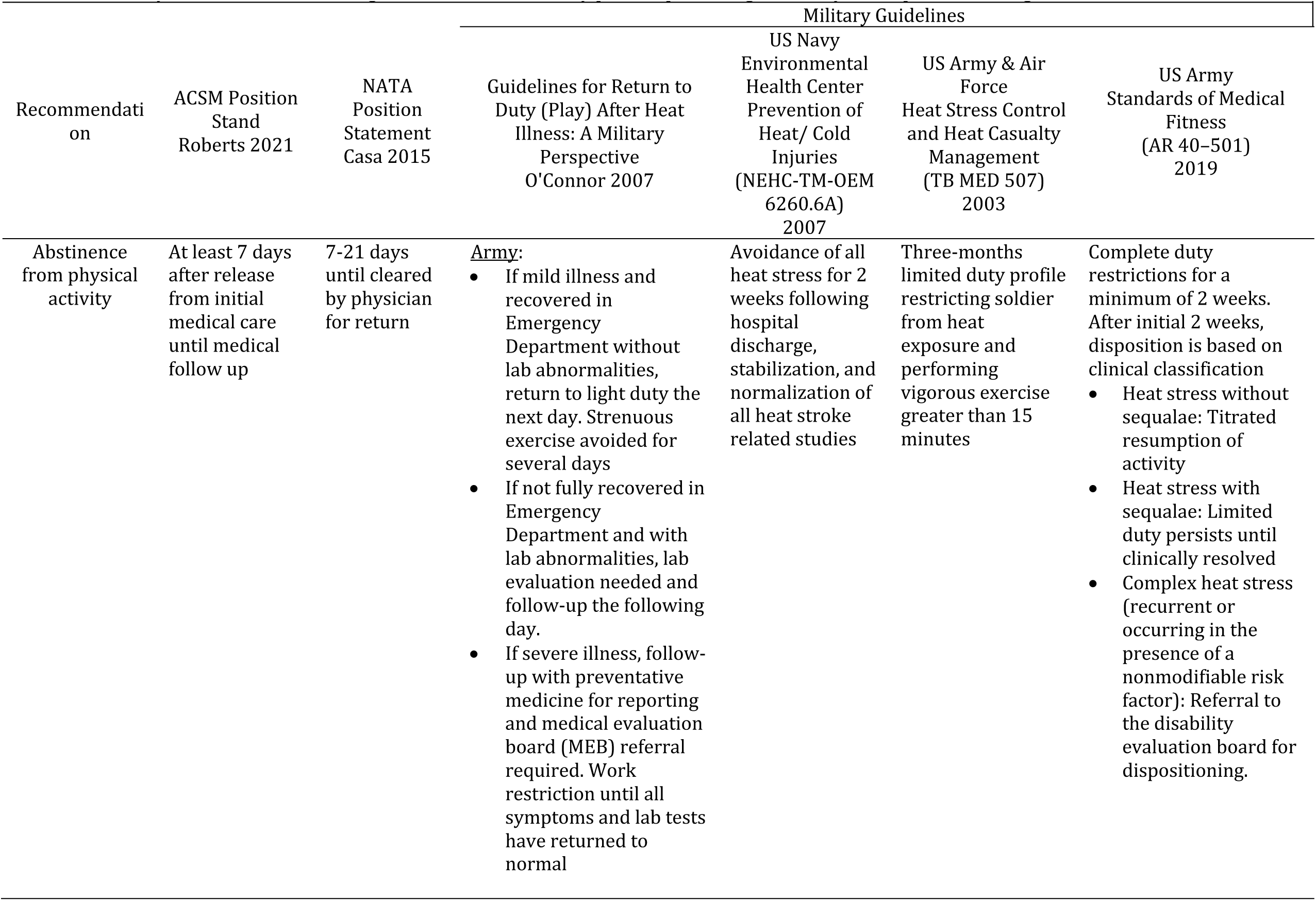
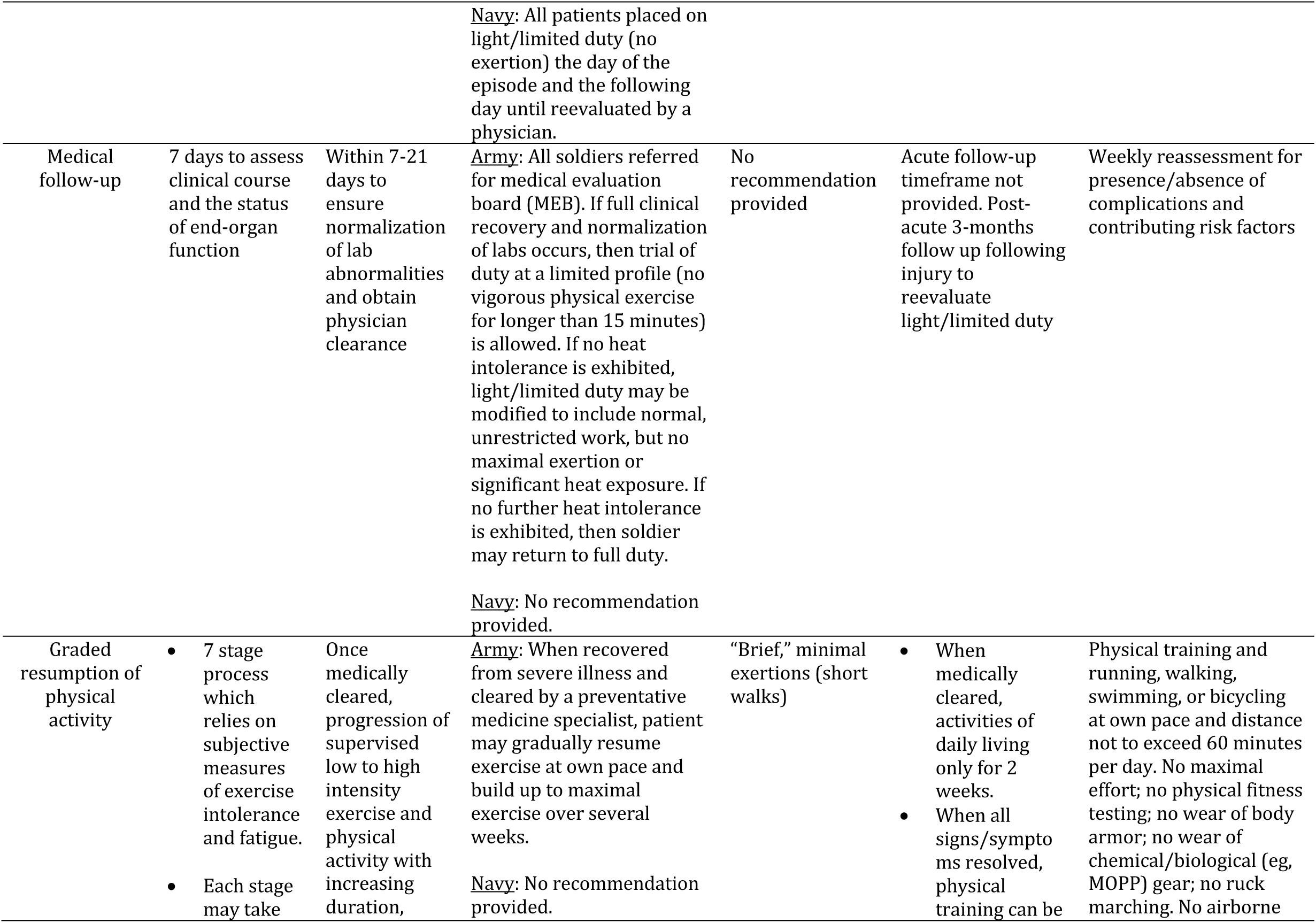
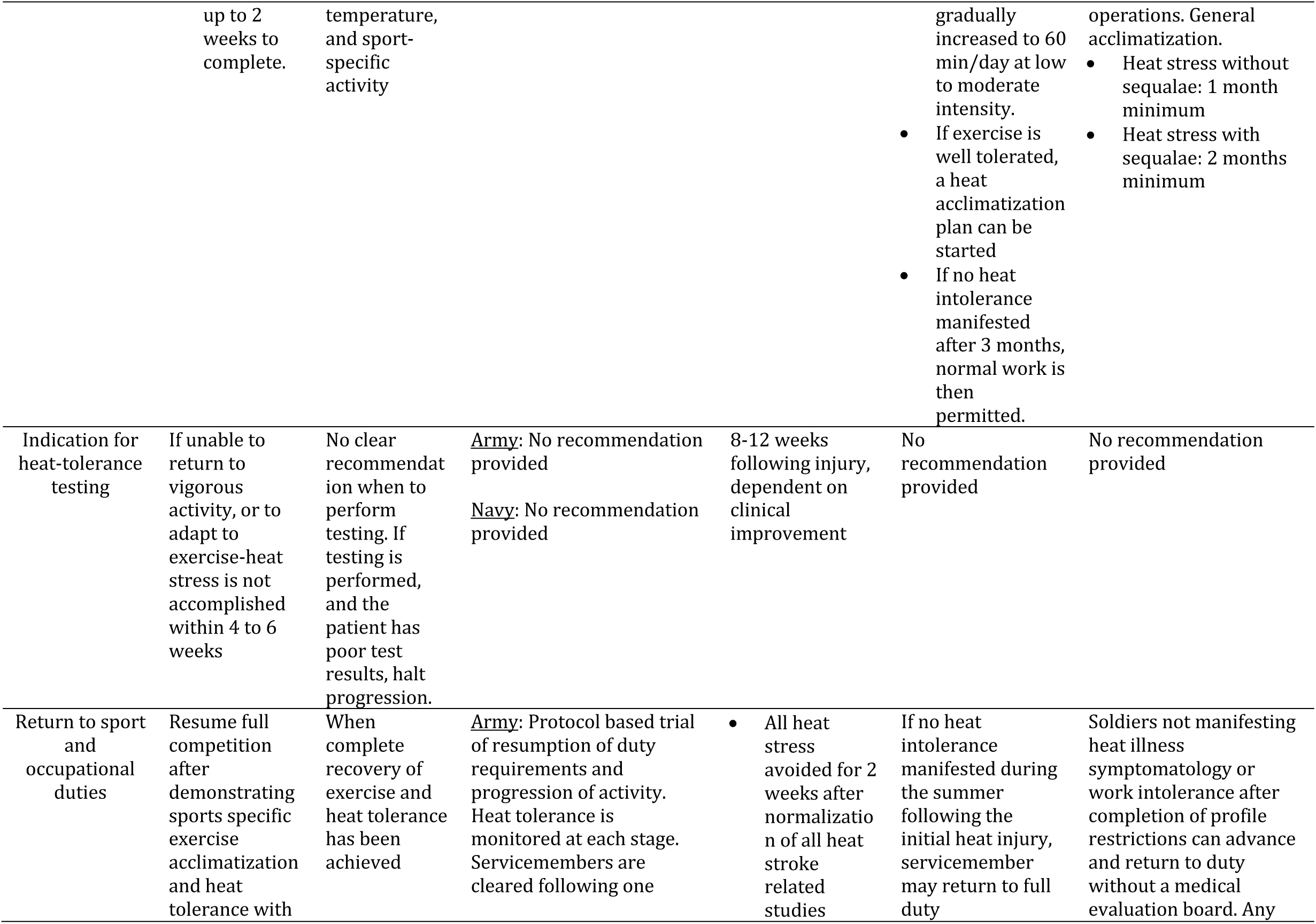
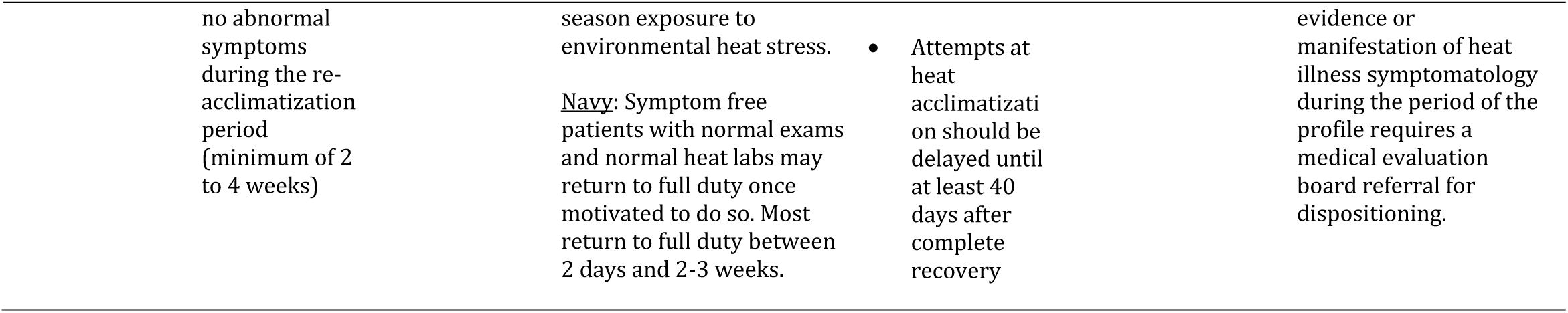
Summary of the included clinical guidelines and US military policies pertaining to activity resumption following exertional heat illness.

### Risk of Bias Assessment

Figure 2. displays the methodological quality of the included studies scored using the six domains covered by the AGREE 2. Both statements had the greatest scores (>80%) in the domains pertaining to scope and purpose, clarity of presentation, and applicability.^9,26^ In domain 2 (stakeholder involvement), both statements scored in the 50-61% range, with the lowest quality observed in the areas pertaining to rigor of development (31-32%) and editorial independence (4-22%).^9,26^

### Study Findings

#### Abstinence From Activity

Both the ACSM consensus statement^26^ and the NATA position statement^9^ recommended abstinence from exercise or physical activity for at least seven days after release from initial medical care. The NATA consensus statement also specified an upper range of 21 days of abstinence until cleared by a physician for return.^9^ Among the military policies, the Navy (NEHC-TM-OEM 6260.6A)^21^ advocated for avoidance of all heat stress for 2 weeks following hospital discharge, stabilization, and normalization of all heat stroke related studies; the Army (AR 40–501)^11^ required a minimum of 2-weeks of abstinence regardless of severity, with continued refrain in individuals with clinical sequalae; and the Army-Air Force (TB MED 507/AFPAM 48-152)^30^ recommended a more conservative 3-month limited duty status restricting soldiers from heat exposure and performing vigorous exercise greater than 15 minutes. O’Connor et al.^23^ also reported finding heterogeneity of recommendations among the service policies. In this report, the authors found that the Army guidelines for return to duty were dependent on severity of illness, patient symptoms, and laboratory values; with the Navy providing specific guidance for cessation the day of illness and the day following until medical reevaluation and clearance for return.^23^

**Figure 2.**
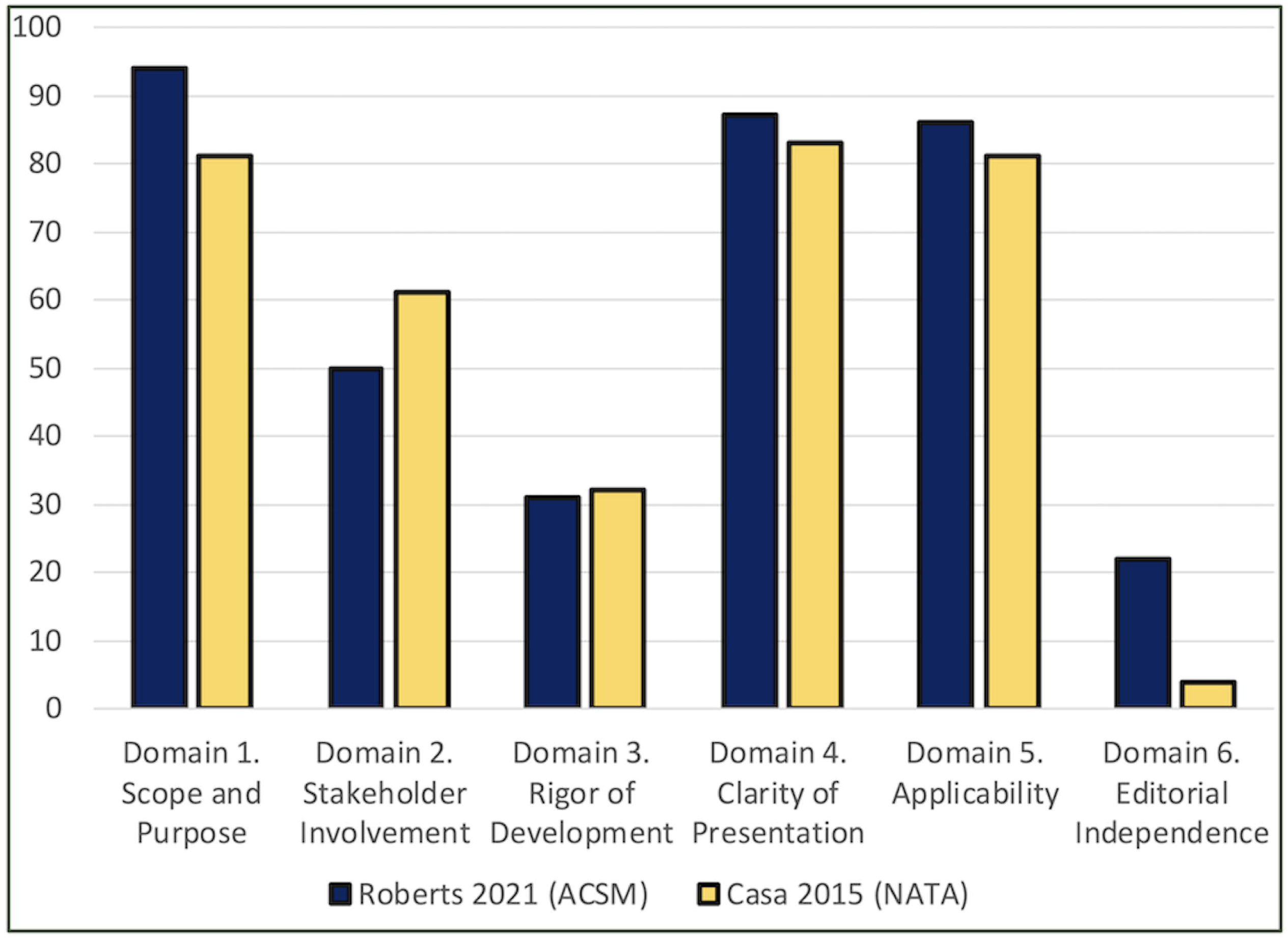
Assessment of methodological quality of included guidelines.

#### Medical Follow-Up

Similar to the recommendations on abstinence, both ACSM and NATA guidelines recommended medical follow-up in 7 days to assess the course of heat illness and monitor organ function,^9,26^ with the NATA statement recommending an upper limit of 21 days for follow up.^9^ In the review of military policy, the Navy policy (NEHC-TM-OEM 6260.6A) did not provide a specific recommendation regarding timeframe for medical follow-up. Current Army-Air Force regulations (TB MED 507/AFPAM 48-152) recommend following up after 3-months to reevaluate light/limited duty.^30^ In the more contemporary Army regulation (AR 40–501), follow up in 7 days followed by weekly reassessments of clinical complications and contributing risk factors was recommended.^11^ In the 2007 brief report, both Army and Air Force policies guided referral of personnel to the medical evaluation board to determine duty status, with follow-up driven based on clinical improvements in laboratory studies and reported symptoms.^23^ The Navy policies in 2007 did not provide specific recommendations pertaining to timeframes for medical follow-up.^23^

#### Graded Resumption of Exercise or Physical Activity

The ACSM guidelines recommend a progressive return to activity that is incrementally reintroduced in a seven-stage process, with progression dependent on subjective reports of exercise intolerance and fatigue during activity.^26^ The NATA guidelines do not specify a protocol for return to activity, but recommends **clinician**-supervised progression from low to high intensity activity once medically cleared.^9^ The Navy policy (NEHC-TM-OEM 6260.6A) similarly recommends a symptom-based return to activity, starting with resumption of low level activity (eg. “brief, minimal exertions” such as short walks) once patients are symptom free, have normal physical examination and laboratory findings, and once motivated to do so.^21^ The Army-Air Force policy (TB MED 507/ AFPAM 48-152) recommends clearance from a preventative medicine specialist following recovery from severe illness prior to resumption of activity.^30^ While the Navy and Army-Air Force policies recommend gradual progression of activity at the service member’s own pace over several weeks once recovered from severe illness and cleared medically, there is no specific guidance provided.^23^ In the more contemporary Army regulation (AR 40–501), a graded resumption of occupational and fitness requirements and re-acclimatization is prescribed based on signs and symptoms of tolerance.^11^

#### Indication for Heat Tolerance Testing

Heat tolerance testing, which consists of assessment of thermoregulation during controlled and monitored physical activity in a hyperthermic environment, was first developed by the Israeli Defense Forces over 30 years ago.^20^ Further details regarding the heat tolerance test (HTT) will be discussed in later sections. The ACSM recommendations suggest that heat tolerance testing be performed for individuals who are unable to return to vigorous activity or adapt to exercise-related heat stress within four to six weeks.^26^ While the NATA statement does not provide clear recommendations for indication or timing or heat tolerance testing, it does specify that physical activity be halted in athletes that do not adequately thermoregulate during testing. None of the reviewed military policies provided **specific** guidance pertaining to this clinical assessment, a finding that was similarly observed in the brief report by O’Connor et al.^23^

#### Return to sport and occupational duties

The ACSM guidelines detail criteria for return to full competition that includes sports-specific exercise acclimatization and heat tolerance with no abnormal symptoms during re-acclimatization period, a period that last for least two to four weeks following illness.^26^ The NATA recommendations are similar and state that clearance may proceed when there is complete recovery of exercise and heat tolerance.^9^ The Navy guidance for return to duty (NEHC-TM-OEM 6260.6A) is dependent on patient signs and symptoms and includes specific criteria that include heat stress avoidance for two weeks after return to normal heat stroke related studies and delay of heat acclimatization until at least 40 days following complete recovery.^21^ The guidance provided in the Army-Air Force policy (TB MED 507/AFPAM 48-152) specifies return to full duty is permissible if there is no heat intolerance manifested during a progressive return to duty and physical training requirements, with full clearance provided the season following exposure to environmental heat stress.^30^ The contemporary Army regulation (AR 40– 501) similarly uses the absence of symptomatology and work intolerance for disposition back to full duty.^11^

### Results of Syntheses

There was convergence between four of the five recommendations provided in the recent ACSM^26^ and NATA^9^ guidelines. Specifically, there was consistency in recommendations pertaining to abstinence of exercises and physical activity timeframes (at least 7 days); medical follow-up in 7 days to assess symptoms, physical signs, and laboratory assessments indicative of organ impairment; graded resumption of physical activity dependent on patient tolerance; and clearance for resumption of all activity when signs and symptoms indicate acclimatization and functional thermoregulation. There was only lack of consistency pertaining to the indication and timeframe for heat tolerance testing,

While there was consistency between the Navy (NEHC-TM-OEM 6260.6A), the Army regulation (AR 40–501), and Army-Air Force (TB MED 507/AFPAM 48-152) policies pertaining to the necessity of abstaining from activity following EHI; monitoring physical signs, laboratory findings, and symptoms; progressive return to activity once signs and symptoms resolve; and trial by training, there is divergence in the details of the recommendations. Specifically, there was substantial heterogeneity observed pertaining to timelines of recovery, with the recommendations provided in the Army-Air Force regulation more specific and conservative than the Navy policy.

When the military policies published in 2003 (TB MED 507/AFPAM 48-152) and 2007 (NEHC-TM-OEM 6260.6A) were contrasted to the contemporary clinical guidelines promulgated by the ACSM^26^ and the NATA,^9^ incongruencies in many of the criteria were noted. The Army regulation (AR 40–501) had the greatest alignment with the professional guidelines that were published around the same time.

## DISCUSSION

The primary finding of this study was that among the multiple medical specialties that manage patients with EHI, only two professional societal guidelines provided clinical recommendations pertaining to return to function. Of these guidelines, there was consistency between recommendations that addressed abstinence from activity; medical follow-up; graded resumption of activity; and return to function. Pertaining to military policy, contemporary regulations published in recent years reflected the best evidence provided in the professional guidelines. The greatest incongruency was noted in older military policies, a likely function of changing practice patterns over the past 15 years.

One of the more surprising findings of this study was that among the sports medicine, family medicine, emergency medicine, occupational medicine, and wilderness medicine disciplines, only two sports medicine societies promulgated specific guidance germane to this topic. While certain disciplines, such as emergency medicine, only manage the patient during the most acute stages of illness and in facilities capable of providing higher levels of care, other disciplines are responsible for patient management throughout the full course of illness and in all possible clinical environments. While the astute clinician will search for evidence among multiple databases and resources within and external to their primary specialty, cooperation between the medical specialty organizations during guideline development may help to improve care through increased diversity of perspective, increase synchrony of recommendations, and facilitate greater dissemination via promulgation through the different societies’ memberships.

The dearth of recommendations from multiple professional societies and inconsistencies between societal guidelines may cause confusion for return to activity after EHI, a sentiment shared by sports medicine providers in Australia.^12^ Sports organizations and military leadership often depend solely on professional societies for guidelines when designing their own policies and regulations. Without consistent and congruent recommendations from the various professional societies, there is a potential risk for organizational interests (e.g. earlier return to sport prior to full recovery for organizational gain) to be prioritized over the individual athlete’s safety. Moreover, there is increased onus on the professional societies to continually update guidelines with the most current evidence available, especially with expectations of exclusive sports organizational use.^12^

Mission execution and personnel readiness are pivotal to the goals of the military. Standardization of protocols and guidelines will help to ensure that the best quality of care is provided across the different branches of the military, while reducing unneeded duplication and waste of resources. Clinical practice is best guided by scientific evidence (to include professional society recommendations), clinical experience, and the individual needs of the patient and organization. However, each branch of the military differs in its exposure of EHI and interpretations of existing guidelines. An increasing number of female tactical athletes now serve in roles that were previously closed to them, a standard that was in effect when many of the existing military EHI policies were published. Since sex has been identified as a non-modifiable intrinsic risk factor for EHI,^1^ it behooves the military to provide updated guidelines with consideration of changes in force composition. Now that the US military health system is aligned under common leadership in the Defense Health Agency with a focus on high reliability practices and standardizing operations across service-branches, the development of updated, consistent clinical pathways among the military service branches is warranted. The recommendations provided by the ACSM^26^ and NATA^9^ pertaining to abstinence of activity, medical follow-up, titrated and clinician-supervised resumption of activity, and progressive environmental exposure to occupational duties following EHI can be readily adopted into military medical policy. Collaboration amongst the service branches will be needed to formulate evidence-based guidelines that can be implemented throughout the US Department of Defense.

Development of military policy that provides sound evidence for clinical management of EHI, while equally considering clinician experience, patient preference, and command objectives in decision-making is essential and integrates all the tenets of evidence-based practice.^27^ There is substantial heterogeneity regarding physical requirements and environments in which a tactical athlete functions. Factors such as service branch, military occupation, and the mode, tempo, and location of military operations are substantive when managing this population. Furthermore, variations in medical training and clinical experience require individualized clinician interpretation and utilization of policy statements for guidance. Striking a balance between clinical direction and protection of clinician autonomy will be essential in the provision of updated clinical pathways and military health policies concerning EHI. Specifically, guidelines that are not dogmatic, prescriptive, and are agnostic to service branch and occupation are recommended, as this will ensure the managing clinician making nuanced clinical decisions based on the individual needs and circumstances of the patient.

Heat tolerance testing is regularly used by the Israeli military as a clinical correlate in the diagnosis of heat intolerance and as criteria for return to function in tactical-athletes.^20^ The HTT protocol was developed to assess thermoregulation in Israeli military members, aged 17 to 30, 6 to 8 weeks following an EHI. The test consists of 120 minutes of treadmill walking at 5 km/h at a 2% grade in 40º C and 40% relative humidity.^19,20^ For safety considerations, testing is aborted if rectal temperature reaches 39° C, heart rate rises above 180 beats/minute, or if the participant develops symptoms of EHI.^20^ Interpretation of results to determine heat tolerance depends on the rectal temperature and the heart rate. Specifically, individuals are classified as heat intolerant if the in-test rectal temperature is greater than 38.5° C, the heart rate rises above 150 beats per minute during the test, or if the change in rectal temperature between 60 and 120 minutes is greater than 0.45.^28^ In addition, an individual is classified as potentially or borderline heat intolerant if the in-test rectal temperature is greater than 38.2° C, the heart rate is between 120 to 150 beats per minute during the test, or if the change in rectal temperature between 60 and 120 minutes is between 0.25° and 0.45° C.^28^ Heat tolerance is achieved if the in-test rectal temperature is less than 38.2° C, in-test heart rate remains less than 120 beats per minute, and the change in rectal temperature between 60 and 120 minutes is less than 0.25.^28^ Typically, the HTT is performed 6 to 8 weeks after EHI and if found to be heat intolerant, they are retested in 3 months.^19,20^ Continued heat intolerance at that time prompts changes in responsibilities and environments within the military.

In contrast, only US Navy policy mentioned indication of heat tolerance testing when making clinical decisions pertaining to disposition following EHI.^21^ Heat tolerance testing is typically indicated when a patient with EHI cannot effectively thermoregulate during physical requirements, especially when exposed to environmental heat and humidity over time.^19,20^ While a positive heat tolerance test is indicative of heat intolerance, this is but one clinical correlate to be used in conjunction with a detailed history, reported symptomatology, physical examination, and other objective measures of potential body system impairment and functional activity. It is unclear if heat tolerance testing improves diagnostic and prognostic accuracy over a comprehensive medical evaluation and more readily available laboratory tests in tactical-athletes with EHI, although there has been evidence correlating heat intolerance as defined by HTT to low cardiorespiratory fitness^16^ and heat tolerance as defined by HTT to significant recurrent EHI risk reduction.^28^ This question warrants further investigation.

The adoption of heat tolerance testing as a standard for return to play or duty has substantial challenges. While the HTT protocol developed by the Israeli military is the most widely used method assessment of thermoregulation, there are alternative protocols that have been developed for military, athletic, occupational medicine, and research applications.^6^ The validity of HTT pertaining to EHI case specificity, individual factors such as heat acclimatization, heat intolerance characteristics, and HTT timing have yet to be established.^6^ Furthermore, heat tolerance testing facilities are not readily accessible, requires the patient to travel to limited specialized testing facilities, and has considerable costs for the equipment and personnel required to operate and interpret results. With the advent of readily available worn or ingestible thermistor sensors, monitoring thermoregulation during function in the execution of sport and duty requirements may be a feasible alternative to traditional testing. While general medical monitoring can be provided locally for patient safety, technology allows for consultation by experts in thermoregulation that can be performed remotely and in real-time. The feasibility and clinical utility of newer technology in assessment of tactical athletes with EHI will require further study.

## Limitations

There are limitations to this study. This systematic review included studies that were only CPGs, consensus statements, position statements, and practice alerts. While it is plausible that practice recommendations included in other study types were omitted due to this delimitation, a boon of this decision was this highlighted an identified gap in the literature for these study types. This study also assessed guidelines specifically written for the clinical management of EHI. It is possible that more generic sport medicine or occupational guidelines that address return to physical activity following illness were not included in this study. Due to the unique presentation and clinical requirements of EHI, this decision was purposefully made *a priori* while acknowledging the potential drawbacks.

## CONCLUSIONS

Despite the multiple medical specialties that manage patients with EHI, only two professional societal guidelines provided clinical recommendations pertaining to return to function during recovery. There was consistency between recommendations that addressed abstinence from activity; medical follow-up; graded resumption of activity; and return to function. Contemporary military regulations published in recent years reflected the best evidence provided in the professional guidelines. The greatest incongruency was noted in older military policies, a likely function of changing practice patterns over the past 15 years. This systematic review highlights the need for consistent recommendation across all branches of the military when it comes to returning servicemembers to duty after EHI.

## Supporting information

Supplemental Table 1. Search strategy

Supplemental Table 2. Characteristics of included guidelines

## Data Availability

All data produced in the present work are contained in the manuscript.

## Acknowledgement

We greatly appreciate the assistance of Simona Konecna and Aileen Y Chang, PhD for their assistance in developing the search strategy and executing the preliminary search.

## List of Tables

**Supplemental Table 1**. Search strategy.

**Supplemental Table 2**. Characteristics of included guidelines

