## Supplemental Table 1. Search strategy for "Consistency and applicability of return to activity guidelines in tactical-athletes with exertional heat illness. A systematic review"

**Ovid MEDLINE search strategy**:
Ovid MEDLINE(R) ALL <1946 to January 28, 2022>
Search performed 31 JAN 2022

1 exp Heat stress disorders/ 6444
2 exp Heat exhaustion/ 1420
3 exp Heat stroke/ 1516
4 *Hot Temperature/ae [Adverse Effects] 4451
5 *body temperature/ or *body temperature regulation/ or *sweating/ 33909
6 exp Hyperthermia/ or Hyperthermia.mp. 38704
7 adverse effects.fs. 1872530
8 (5 or 6) and 7 6383
9 ((heat adj1 stress) or (heat adj1 exhaust$) or (heat adj1 stroke$) or (heat adj1 cramp$) or (heat adj1 syncope$) or (heat adj1 injur$) or (heat adj1 ill$) or (heat adj1 tolerance) or (heat adj1 intolerance) or (heat adj1 strain$) or (heat adj1 exposure) or (heat adj1 acclim$) or (heat adj1 wave$) or (exertional adj1 heat adj1 stroke) or "heatwave$" or "heatstroke$").mp. 26088
10 ((hot adj1 temperature) or (body adj1 temperature) or sweating or overheat$ or hyperthermia or (hot adj1 weather)).ti,ab. 66255
11 1 or 2 or 3 or 4 or 8 or 9 or 10 93302
12 Guideline/ or Practice Guideline/ 36743
13 "guidelines as topic"/ 41908
14 (guideline$ or (practice adj1 guideline$) or (position adj1 statement$)).mp. 542279
15 12 or 13 or 14 542279
16 11 and 15 1298
17 limit 16 to english language 1183

**Web of Science search strategy**:
Databases: Web of Science Core Collection – SCI-Expanded, SSCI; date searched: 31 JAN 2022

| Set | | # records |
|---|---|---|
| #1 | TS = ("heat stress disorder*") | 60 |
| #2 | TS = ("heat exhaust*") | 431 |
| #3 | TS = ("heat stroke") | 1784 |
| #4 | TS = (hot temperature adverse effects) | 619 |
| #5 | TS = ("body temperature" or "body temperature regulation" or "sweating") | 40,583 |
| #6 | TS = (hyperthermia) | 38,253 |
| #7 | TS = ("adverse effects") | 141,732 |
| #8 | (#5 OR #6) AND #7 | 910 |
| #9 | TS = ((heat NEAR/1 stress) or (heat NEAR/1 exhaust*) or (heat NEAR/1 stroke$) or (heat NEAR/1 cramp*) or (heat NEAR/1 syncope$) or (heat NEAR/1 injur*) or (heat NEAR/1 ill*) or (heat NEAR/1 tolerance) or (heat NEAR/1 intolerance) or (heat NEAR/1 strain$) or (heat NEAR/1 exposure) or (heat NEAR/1 acclim*) or (heat NEAR/1 wave$) or (exertional NEAR/1 heat NEAR/1 stroke) or "heatwave*" or "heatstroke*") | 56,444 |

| | | |
|---|---|---|
| #10 | TI = ((hot NEAR/1 temperature) or (body NEAR/1 temperature) or sweating or overheat* or hyperthermia or (hot NEAR/1 weather)) OR AB = ((hot NEAR/1 temperature) or (body NEAR/1 temperature) or sweating or overheat* or hyperthermia or (hot NEAR/1 weather)) | 94,354 |
| #11 | (#1 OR #2 OR #3 OR #4 OR #8 OR #9 OR #10) | 145,834 |
| #12 | TS = (guideline$ or "practice guideline$" or (practice NEAR/1 guideline$) or (position NEAR/1 statement$)) | 558,264 |
| #13 | #11 AND #12 | 2,045 |
| #14 | limit 13 to English language | 1,931 |

CINAHL Complete search strategy (via EBSCOHost platform)
Search performed 31 JAN 2022

| Set | | # records |
|---|---|---|
| #1 | (MH "Heat Stress Disorders+") | 2,458 |
| #2 | "hot temperature" | 20 |
| #3 | (MH "Body Temperature+") OR (MH "Body Temperature Regulation+") OR (MH "Thermogenesis+") | 11,921 |
| #4 | "hyperthermia" | 9,473 |
| #5 | "adverse effects" | 513,217 |
| #6 | S5 AND (S3 OR S4) | 3,086 |
| #7 | ((heat W1 stress) or (heat W1 exhaust*) or (heat W1 stroke*) or (heat W1 cramp*) or (heat W1 syncope*) or (heat W1 injur*) or (heat W1 ill*) or (heat W1 tolerance) or (heat W1 intolerance) or (heat W1 strain*) or (heat W1 exposure) or (heat W1 acclim*) or (heat W1 wave*) or (exertional W1 heat W1 stroke) or "heatwave$" or "heatstroke$") | 4,152 |
| #8 | TX((hot W1 temperature) or (body W1 temperature) or sweating or overheat* or hyperthermia or (hot W1 weather)) | 42,140 |
| #9 | S1 OR S2 or S6 OR S7 OR S8 | 45,722 |
| #10 | (MH "Guideline Adherence") OR (MH "Practice Guidelines") | 94,741 |
| #11 | (guideline* or (practice W1 guideline*) or (position W1 statement*)) | 226,351 |
| #12 | S10 or S11 | 226,351 |
| #13 | S9 AND S12 | 1,743 |
| #14 | Limit 13 to English | 1,704 |
| #15 | Remove duplicates in EndNote (Author, Year, Title, Journal) | 1,597 |
