## Supplemental Table 2. Characteristics of included guidelines for "Consistency and applicability of return to activity guidelines in tactical-athletes with exertional heat illness. A systematic review"

| Author | Year Published | Guideline Type | Objective | Questions | Population | Search Methods | Inclusion Criteria | Exclusion Criteria | Recommendatrion development process | Recommended criteria for return to sport/occupation |
|---|---|---|---|---|---|---|---|---|---|---|
| Casa, DeMartini, Bergeron, Csillan, Eichner, Lopez, Ferrara, Miller, O'Connor, Sawka, Yeargin (NATA Position Statement: Exertional Heat Illness 2015) | 2015 | Position Statement | Present best-practice recommendations for prevention, recognition, and treatment of exertional heat illness | Best practice recommendations for managing heat related illness | Athletes | No mention | No mention | No mention | Strength of recommendations graded using SORT criteria | Exercise-associated muscle cramps (EAMC's) or heat syncope: Monitor until signs and symptoms are no longer present<br>Heat exhaustion: same day return is not recommended<br>Exertional Heat Stroke: Patients cooled effectively may be able to resume modified activity within 1 month with a physician's clearance. Most guidelines suggest patients be asymptomatic with normal blood-work results (renal, hepatic, electrolytes, muscle enzyme levels) before gradual return to activity is initiated. Few evidence based strategies exist, so must rely on clinical signs, responses to heat-tolerance tests, responses to gradually increasing exercise demands, and ability to acclimatize to heat.<br>In all cases, prior to beginning progression of activity of increasing intensity and duration, patient much have complete a 7 to 21 day rest period, demonstrated normal blood work, and obtained physician clearance. Rectal temperature and heart rate should be monitored<br>Heat tolerance testing: As research has advanced, role of testing has gained favor. Patient who has a poor test result should not increase activity at that point. The significance of a normal test result and it's relationship with clearance to return to play still needs to refined and evaluated |
| Roberts, Armstrong, Sawka, Yeargin, Heled, O'Connor (ACSM Position Statement 2021) | 2021 | Consensus Statement | Identify evidence based strategies to reduce morbidity and mortality of exertional heat illness, including introducing a staged return to activity (RTA) for athletes recovering from an EHS event. | What are the most uo to date, evidence base strategies to reduce morbidity and mortality of exertional heat illness, including introducing a staged return to activity (RTA) for athletes recovering from an EHS event. | Athletes & General population | No mention | No mention | No mention | Consensus based approach | Evidence-based data to facilitate RTA following EHI or EHS is limited and individual recovery following severe exertional heat illness is highly variable. Research is needed to evaluate the role of a graduated increase in structured physical exercise to assist in RTA decisions.<br><br>Recommendations: Should have full symptom resolution at rest and normal laboratory findings for organs most often affected by EHI or EHS (e.g., liver and kidney) before starting a cautious reintroduction of physical activity and a gradual heat acclimatization program. Assessing the basic hematologic parameters and blood chemistries for normal renal, hepatic, and coagulation normal function will give a baseline at the onset of activity. An evaluation about 1-wk post-incident for a physical examination and laboratory testing or diagnostic imaging (biomarkers) of the affected organs is suggested for EHS follow-up. Following EHI or EHS, a medical evaluation of the patient is completed every week until all symptoms, signs, and abnormal laboratory values have resolved. Activities of daily living are the only "exercise" allowed for 2 wk. When all the symptoms and signs have resolved, physical training can be gradually increased to 60 min·d$^{-1}$ at low to moderate intensity. When this level of exercise is well tolerated, a heat acclimatization plan (gradually increasing the duration and intensity of exercise and heat stress each day) can be started. If no symptoms of heat intolerance or abnormal blood work values are observed, the patient is released to unsupervised sport specific physical activity.<br><br>Stages for returning to activity following EHS: The return protocol relies on subjective measures, and the athlete is advanced to the next stage if there is no evidence of exercise intolerance or fatigue. Each stage is individualized but may require up to 2 weeks to complete in EHS victims. If an athlete is unable to appropriately advance (stages 3 through 6), additional evaluation may be needed to determine the capacity for strenuous exercise in the heat, which may include heat tolerance testing (HTT) to assess current heat tolerance.<br>Stage 1: Early Medical Recovery Physician-guided Organ system recovery; Activities of daily living for 1 to 2 week; Gradual increase in home activities without fatigue<br>Stage 2: Mid Medical Recovery Physician Guided Sustain minimal aerobic fitness and develop confidence; Self-paced comfortable walk in low heat stress conditions (e.g., an air-conditioned gymnasium); 20–60 min at maximal intensity of HR < 100 or <50% Age-Adjusted maximal HR<br>Stage 3: Early Exercise Adaptation Athletic trainer guided with physician; Gradually improve aerobic exercise capability; Walk at 3.5 mph in low heat stress conditions; 60 min at HR < 140 bpm or <70% of age adjusted maximal HR<br>Stage 4: Mid Exercise Adaptation Athletic Trainer Guided with Physician Gradually improve aerobic exercise capability and fitness; Walk and run in low heat stress conditions; 60 min of progressively increasing run to walk ratio until constant run for 60 min<br>Stage 5: Heat Acclimatization Athletic Trainer Guided Gradually improve heat acclimation status; Run in ambient warm or hot conditions; 60 min of progressively increasing run until constant run for 60 min<br>Stage 6: Sports-Specific Acclimatization/Training Athletic trainer and/or coach guided. Improve sport specific heat acclimation and fitness; Participate in practice in ambient conditions; Initially participate in sports specific drills with sports specific equipment then progress to training and scrimmage<br>Stage 7: Full Return to Sport Athletic trainer monitors during warm-up and game; Normal game or competition participation in ambient conditions<br><br>RTA Considerations: A detailed history and physical examination including unique intrinsic and extrinsic risk factors, the timing of treatment, and the rate of cooling must be considered it the RTA decision. The athlete should refrain from exercise for at least 7 d after release from initial medical care, at which time the clinician will address the clinical course of the heat stroke incident and carefully assess the status of end-organ function (neurocognitive, renal, hepatic, muscle, hematologic as clinically indicated). The clinician should carefully address any intrinsic and extrinsic risk factors associated with the EHS event. When medically eligible for RTA/RTP based upon the return of normal end organ function, an individual can begin exercise in a cool environment and gradually increase the duration, intensity, and heat exposure over 2 to 4 weeks to initiate environmental acclimatization, improve fitness, and demonstrate heat tolerance. If return to vigorous activity and evidence of the patient's ability to adapt to exercise-heat stress over several days is not accomplished within 4 to 6 weeks, consider referral to a physician with experience in heat-related disorders for further evaluation that may include HTT in a controlled setting. The timing for full RTP is highly variable based upon the inter-individual severity and recovery of each EHS event. In general, an athlete may be allowed to resume full competition after demonstrating sports specific exercise acclimatization and heat tolerance with no abnormal symptoms during the re-acclimatization period; this process normally requires a minimum of 2 to 4 weeks. |
